# Computerized analysis of the eye vasculature in a mass dataset of digital fundus images: the example of age, sex and primary open-angle glaucoma

**DOI:** 10.1101/2024.07.21.24310763

**Authors:** Jonathan Fhima, Jan Van Eijgen, Anat Reiner-Benaim, Lennert Beeckmans, Or Abramovich, Ingeborg Stalmans, Joachim A. Behar

## Abstract

**Objective:** To develop and validate an automated end-to-end methodology for analyzing retinal vasculature in large datasets of digital fundus images (DFIs), aiming to assess the influence of demographic and clinical factors on retinal microvasculature.

**Design:** This study employs a retrospective cohort design to achieve its objectives.

**Participants:** The research utilized a substantial dataset consisting of 115,237 digital fundus images obtained from individuals undergoing routine eye examinations. There was no inclusion of a separate control group in this study.

**Methods:** The proposed methodology integrates multiple stages: initial image quality assessment, detection of the optic disc, definition of the region of interest surrounding the optic disc, automated segmentation of retinal arterioles and venules, and the engineering of digital biomarkers representing vasculature characteristics. To analyze the impact of demographic variables (age, sex) and clinical factors (disc size, primary open-angle glaucoma [POAG]), statistical analyses were performed using linear mixed-effects models.

**Main Outcome Measures:** The primary outcomes measured were changes in the retinal vascular geometry. Special attention was given to evaluating the independent effects of age, sex, disc size, and POAG on the newly engineered microvasculature biomarkers.

**Results:** The analysis revealed significant independent similarities in retinal vascular geometry alterations associated with both advanced age and POAG. These findings suggest a potential mechanism of accelerated vascular aging in patients with POAG.

**Conclusions:** This novel methodology allows for the comprehensive and quantitative analysis of retinal vasculature, facilitating the investigation of its correlations with specific diseases. By enabling the reproducible analysis of extensive datasets, this approach provides valuable insights into the state of retinal vascular health and its broader implications for cardiovascular and ocular health. The software developed through this research will be made publicly available upon publication, offering a critical tool for ongoing and future studies in retinal vasculature.

---

The eye is a sensory organ attuned to visible light and a crucial component of the sensory nervous system. Its transparent media serve as a unique, non-invasive window to the vascular system, vulnerable to a variety of diseases. The ocular vasculature can be non-invasively examined using digital fundus images (DFIs). This facilitates unprecedented opportunities to analyze the retinal microvasculature and its potential connection to health disorders. In 1999, Sharrett et al.^1^ identified arterio-venous nicking and arteriole narrowing as pathological findings of hypertension. Their study involved analysis of the central retinal artery equivalent (CRAE) and the central retinal vein equivalent (CRVE) in a cohort of 9,040 patients. In 2006, Witt et al.^2^ described how the decrease in tortuosity in the retinal arterioles was associated with an increased risk of death due to ischemic heart disease (n= 680) in the Beaver Dam Eye Study. In 2009, Sabanayagam et al. reported that individuals with reduced CRAE were more likely to have chronic kidney disease than those with increased CRAE (n=2581)^3^. Regarding primary open-angle glaucoma (POAG), several studies found lower CRAE in POAG patients compared to healthy controls; however, none of these studies included more than several hundred glaucoma cases^4–13^. Similar to CRAE but to a lesser extent, lower CRVE is commonly found in this population^4, 6–13^. In the subsequent decade, fractal dimension of the retinal vascular tree emerged as a risk predictor of cardiovascular disease. Studies conducted between 2008 and 2011 with patient cohorts ranging from 166 to 3,303 participants showed that monofractal dimension could vary in association with age, smoking behavior, blood pressure, diabetic retinopathy, chronic kidney disease, stroke, and mortality due to coronary heart disease^14–19^. In 2021, retinal multifractal dimensions (n= 2,333) were associated with blood pressure and cardiovascular disease^20^. In POAG patients, a lower arteriolar and venular fractal dimension was observed compared to healthy controls, however, these studies are small (n = 87 and n = 61 POAG cases)^4, 21^. In 2022, a study of 1,770 patients suggested that a set of vascular biomarkers could undergo changes in response to hypertension. The study calculated 10 vascular biomarkers (VBMs) in a semi-automated manner using the SIVA software^22^. Recent studies have used deep learning to analyze large-scale datasets^23, 24^, but these models often lack explainability, impeding physiological insights.

Despite growing advances and the diversity of patient cohorts investigated over the years, these studies share common limitations. The semi-automated nature of the computation might introduce some discrepancies in results depending on the grader, and hinder scalability due to the intensive human supervision required for laborious accurate analysis. Furthermore, while CRAE, CRVE and AVR are computed within a specified area of the DFIs, this is not necessarily the case for other biomarkers. Some studies^25, 26^ have used zones A, B, and C, originally devised for CRAE, CRVE, and AVR measurements, to calculate different VBMs. Nevertheless, this method might be prone to bias as these zones are determined based on the size of the optic disc (OD), which can vary due to conditions like POAG^27^.

This research introduces an end-to-end method for analyzing the retinal vasculature in large DFI datasets. Deep learning techniques are employed to automatically extract the optic disc and blood vessels from a DFI, as well as to classify these blood vessels into arterioles (A) and venules (V). Using these automated extractions, we propose a method for standardizing vascular biomarker analysis, based on a fixed-sized region of interest (ROI) around the optic disc. This minimizes the influence of image centering around the OD. Our methods include statistical analysis using linear mixed models to evaluate the effects of demographics or clinical variables on individual microvascular biomarkers. The method is applied to a dataset of 100,000 DFIs for evaluating the effects of age, sex, and POAG on the microvasculature. This novel approach enables quantitative analysis of ocular vasculature and supports the study of its association with specific diseases, facilitating reproducible analysis of large datasets. The resulting software is made available to the community.

## Method

### Study Design and participants

We used the Leuven UZF dataset^28, 29^ which consists of a total of 115,237 DFIs from 13,185 unique participants, captured between 2010 and 2019 in the Glaucoma Clinic of the University Hospitals UZ Leuven in Belgium (IRB No S60649). The DFIs of the UZF dataset are disc-centered and acquired using a Visucam Pro NM camera with 30° FOV (Zeiss) and a resolution of 1444×1444 pixels. Low-quality images, defined as FundusQ-Net less than six were excluded^30^. Each of the DFIs was labeled with no, one or more medical diagnosis codes (ICD10). The codes were reviewed and categorized as Healthy, POAG, or other as defined in section 2.1. Finally, DFIs where the OD could not be segmented or where the ROI was less than 500 px in radius were excluded. This led to a final dataset containing 32,768 DFIs from 4858 unique participants. Fig. 1 summarizes the data selection steps.

**Fig. 1:**
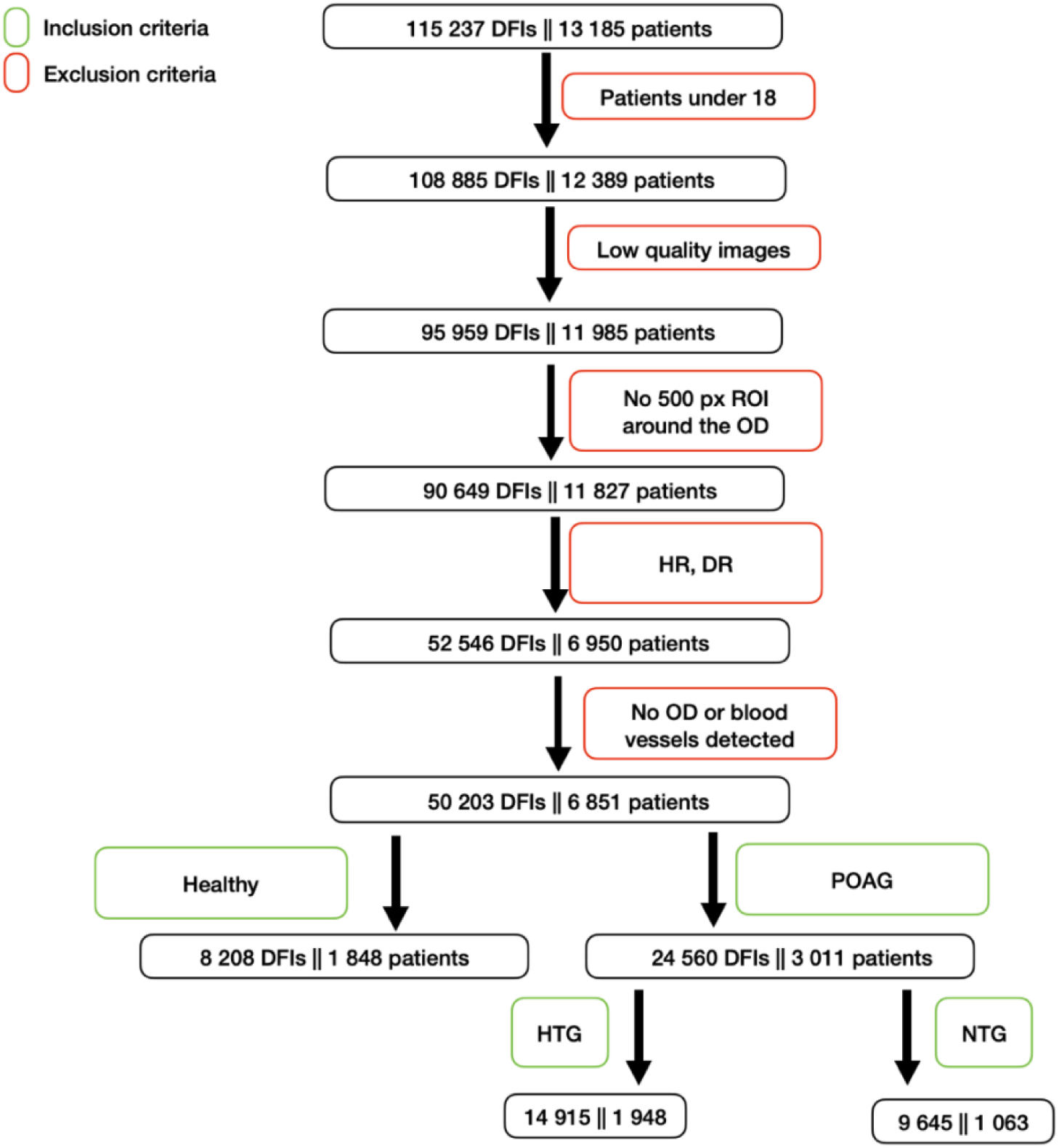
Summary of the dataset elaboration.

Children below 18 years of age and patients with diabetic or hypertensive retinopathy were excluded from the dataset. POAG-labeled eyes, either normal tension glaucoma (NTG) or high-tension glaucoma (HTG) based on the maximal untreated intra-ocular pressure (21 mmHg cut-off), were included until or unless they were diagnosed with other eye diseases, congenital diseases, non-glaucoma surgery or interventions. The absence of disease labels resulted in classification in the healthy group.

### Automated Biomarkers Extraction

LUNet^28^ was used to segment the OD and A/V from the DFI. A ROI with a fixed size of 500 pixels was extracted around the OD (excluding the OD), along with zones A and B, for all DFIs. Subsequently, the CRAE, CRVE, and AVR were calculated using the A/V from zone B. To correct for variability of the zone A and B, the radius of zone A, named disc radius a, was adjusted for. In contrast, other VBMs were computed within the fixed-size ROI for the arterioles and venules (Fig. A2). The overall pipeline is summarized in Fig. 2. DFIs without an ROI of at least 500 pixels around the OD were excluded.

**Fig. 2:**
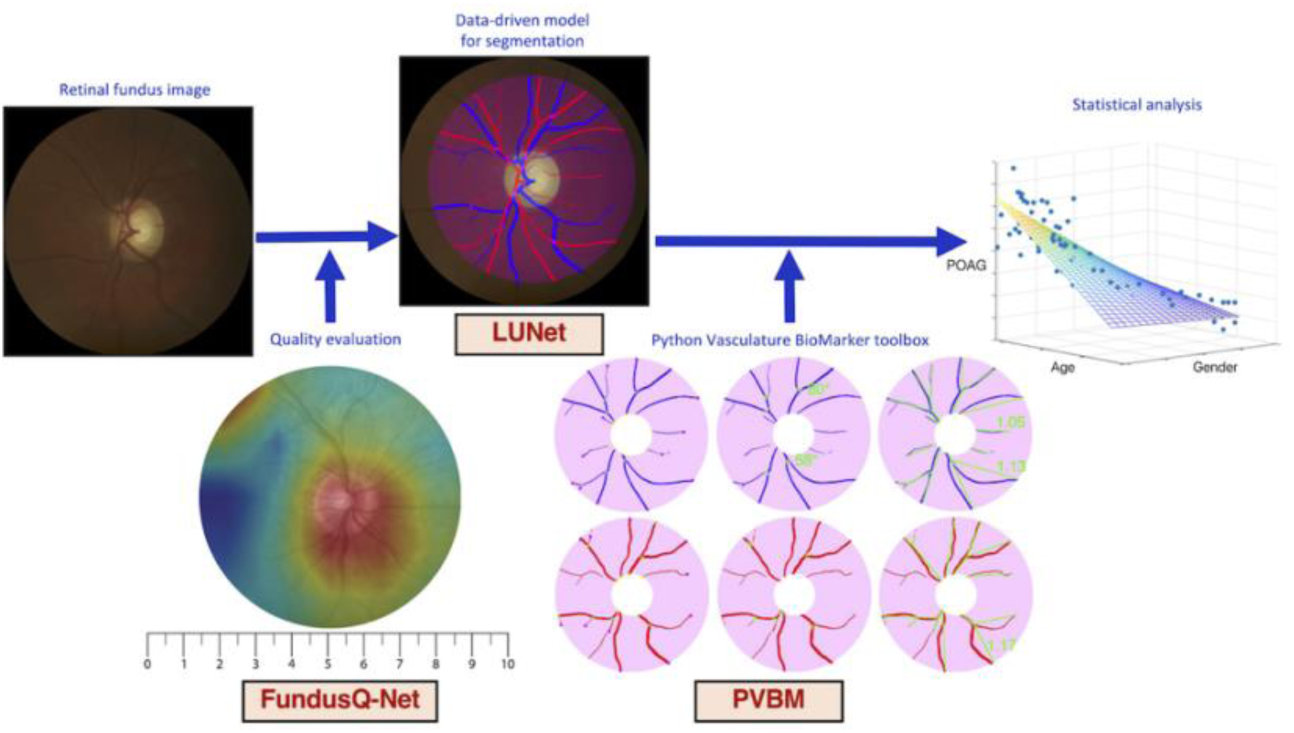
Overview of the experiments. The quality of the digital fundus images (DFI) is first automatically computed using FundusQ-Net^30^. DFI of sufficient quality are processed with LUNet^29^ for arteriole/venule (A/V) and optic disc segmentation. A region of interest is defined around the optic disc to standardize the analysis, and a set of digital vasculature biomarkers is computed using the PVBM toolbox^31^. Finally, a statistical analysis of the vasculature characteristics is performed as a function of demographic and clinical variables.

To define a large-scale vasculature analysis pipeline, it is necessary to segment the A/V and the OD from the DFIs. LUNet^28^ was used to perform a high-resolution A/V segmentation. LUNet was trained to perform A/V segmentation at the same resolution as the DFIs from the UZF dataset. Briefly, LUNet architecture includes a double dilated convolutional block that aims to enhance the receptive field of the model and reduce its parameter count.

The custom loss function emphasizes the continuity of blood vessels. OD segmentation does not require the same fine-grained segmentation as the A/V task. For efficient segmentation of the OD, LUNet was trained on DFIs that were downscaled to a resolution of 256×256 pixels. This downsizing of the DFIs was carried out using bilinear interpolation. Subsequently, the probability map generated by LUNet was upscaled back to the original resolution of the DFIs using bilinear interpolation, resulting in a segmentation map that matched the original dimensions of the DFIs. An example of DFI segmentation can be seen in Fig. A2.

In POAG follow-up, DFIs are often disc-centered. However, this centering is never exact, and this affects the estimation of several VBMs. To standardize the biomarker computation and minimize this effect, an ROI centered on the OD is defined. The ROI radius was set to 500 pixels, which resulted in 95% retention of the segmentation for analysis (see Fig. A1). An example of the ROI extracted is presented in Fig. 3. Finally, the VBM was engineered for A/V separately using the PVBM^31^ toolbox. The set of VBM included is summarized in Table 1. Examples of engineered VBMs inside the ROI can be seen in Fig. 2.

**Fig. 3:**
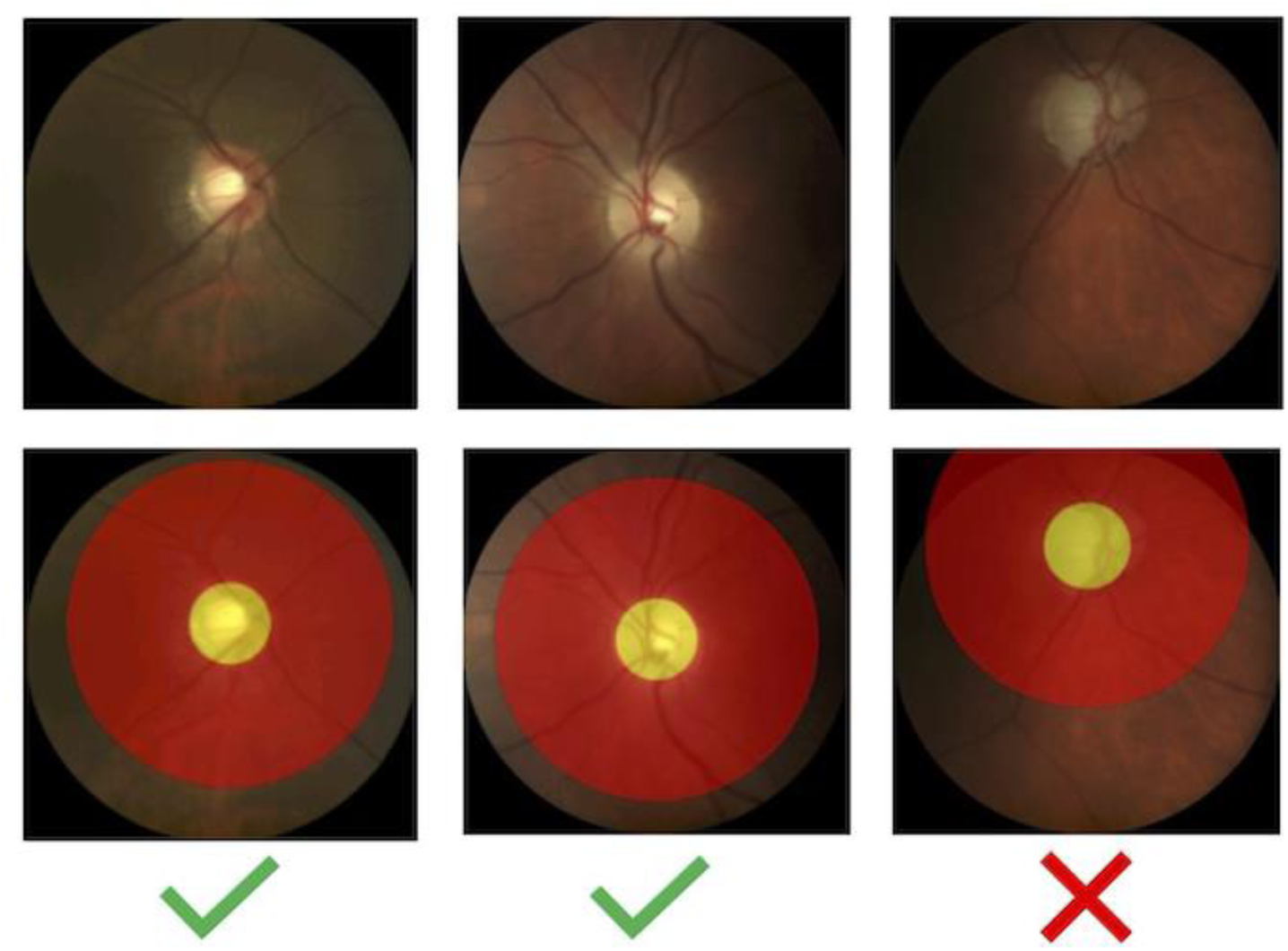
Example of DFIs accepted and discarded based on the availability of the ROI.

**Table 1:** List of digital vasculature biomarkers implemented in PVBM^31^.

| Number | Biomarker | Definition | Unit |
| --- | --- | --- | --- |
| 1 | OVLEN<sup>32</sup> | Overall length | Pixel |
| 2 | AREA<sup>32</sup> | Overall area | Pixel<sup>2</sup> |
| 3 | START<sup>32</sup> | Number of starting points | - |
| 4 | END<sup>32</sup> | Number of endpoints | - |
| 5 | INTER<sup>32</sup> | Number of intersection points | - |
| 6 | TOR<sup>32, 33</sup> | Median tortuosity | % |
| 7 | TI<sup>32</sup> | Tortuosity index | % |
| 8 | BA<sup>32</sup> | Branching angles | °(degree) |
| 9 | D<sub>0</sub><sup>34, 35</sup> | Capacity dimension | - |
| 10 | D<sub>1</sub><sup>34, 35</sup> | Entropy dimension | - |
| 11 | D<sub>2</sub><sup>34, 35</sup> | Correlation dimension | - |
| 12 | SL<sup>36</sup> | Singularity Length | - |
| 13 | CRAE<sub>h</sub><sup>37</sup> | Central retinal artery equivalent using the formulas defined by Hubbard et al. | Pixel |
| 14 | CRVE<sub>h</sub><sup>37</sup> | Central retinal artery equivalent using the formulas defined by Hubbard et al. | Pixel |
| 15 | AVR<sub>h</sub><sup>37</sup> | Arterio-venous ratio using the formulas defined by Hubbard et al. | - |
| 16 | CRAE<sub>k</sub><sup>38</sup> | Central retinal arterial equivalent using the formulas defined by Knudtson et al. | Pixel |
| 17 | CRVE<sub>k</sub><sup>38</sup> | Central retinal venular equivalent using the formulas defined by Knudtson et al. | Pixel |
| 18 | AVR<sub>k</sub><sup>38</sup> | Arterio-venous ratio using the formulas defined by Knudtson et al. | - |
| 19 | AVR<sub>h</sub><sup>38</sup> | Arterio-venous ratio using the formulas defined by Knudtson et al. or Hubbard et al. | - |

### Statistical analysis

Digital VBM levels were compared between Healthy and POAG eyes using the Mann-Whitney U test as all biomarkers were non-normally distributed (Anderson-Darling test). Since the groups did differ in age and sex, a multivariate linear mixed-effects model (LMM) was used to evaluate and correct for the effect of age, sex, disc radius, and POAG on each VBM. The random effects of the LMM additionally account for the dependence between multiple DFIs of the same patient. Based on the central limit theorem, LMM analysis can be validly interpreted. The LMM was trained on the full 32,768 images. The resulting standardized model coefficients (Z statistics) were presented for all VBMs as a heatmap. For the Mann-Whitney U-tests, only the first image of every patient was used, randomly sampled for the left or right eye if both were eligible for analysis. The p-values were adjusted for multiplicity using the Benjamini-Hochberg FDR controlling procedure^39^. The 0.05 level was used for significance.

## Results

After selection, 3010 POAG patients and 1848 healthy controls were included in this experiment. The average age of the POAG cohort (70 years) was higher than that of the control cohort (59 years, p < 0.001), as was the proportion of men (47% vs 40% respectively, p<0.001). The demographics, as well as the proportion of NTG and HTG, can be found in Table 2. Overall, the POAG cohort is characterized by moderate disease. As age, sex and disc radius differ between groups, adjustment for these covariates is necessitated.

**Table 2:** Demographics by diagnosis.

| Demographics Variable | Normal<br>(N=1848) | POAG<br>(N=3010) | POAG Subtypes |  |
| --- | --- | --- | --- | --- |
|  |  |  | NTG (N=1063) | HTG (N=1948) |
| Age (years, mean ± std) | 58.54 ± 16.92 | 69.67 ± 11.39 | 70.13 ± 11.34 | 69.42 ± 11.42 |
| MD (dB, mean ± std) | - | L: -9.01 ± 7.76<br>R: -8.38 ± 7.73 | L: -8.36 ± 7.06<br>R: -7.68 ± 7.06 | L: -9.39 ± 8.12<br>R: -8.78 ± 8.06 |

Table 3 provides summary statistics for the VBMs of the eyes of POAG patients and Healthy participants with a comparison between the groups. All VBMs are lower in POAG patients compared to healthy controls, except for arteriolar and venular singularity length and the Knudtson arterio-venous ratio, after adjusting for multiple testing. The multivariate linear mixed models equally show that the VBMs are influenced by POAG, age, sex, and the disc radius a. These results are presented in Fig. 4. With age all VBMs are uniformly lower except for singularity length and branching angle. Men, when POAG status, age, and disc diagnosis are held constant, have lower arteriolar and venular diameter, area and length, lower arteriolar and venular monofractal and multifractal dimension, lower venular branching angles, and higher arteriolar branching angles. Larger disc size does significantly impact the VBMs, resulting in more starting points and less overall vessel area, which prompts proper statistical adjustment. Although the arteriolar and venular area and length are lower, the CRAE and CRVE tend to be slightly larger.

**Fig. 4:**
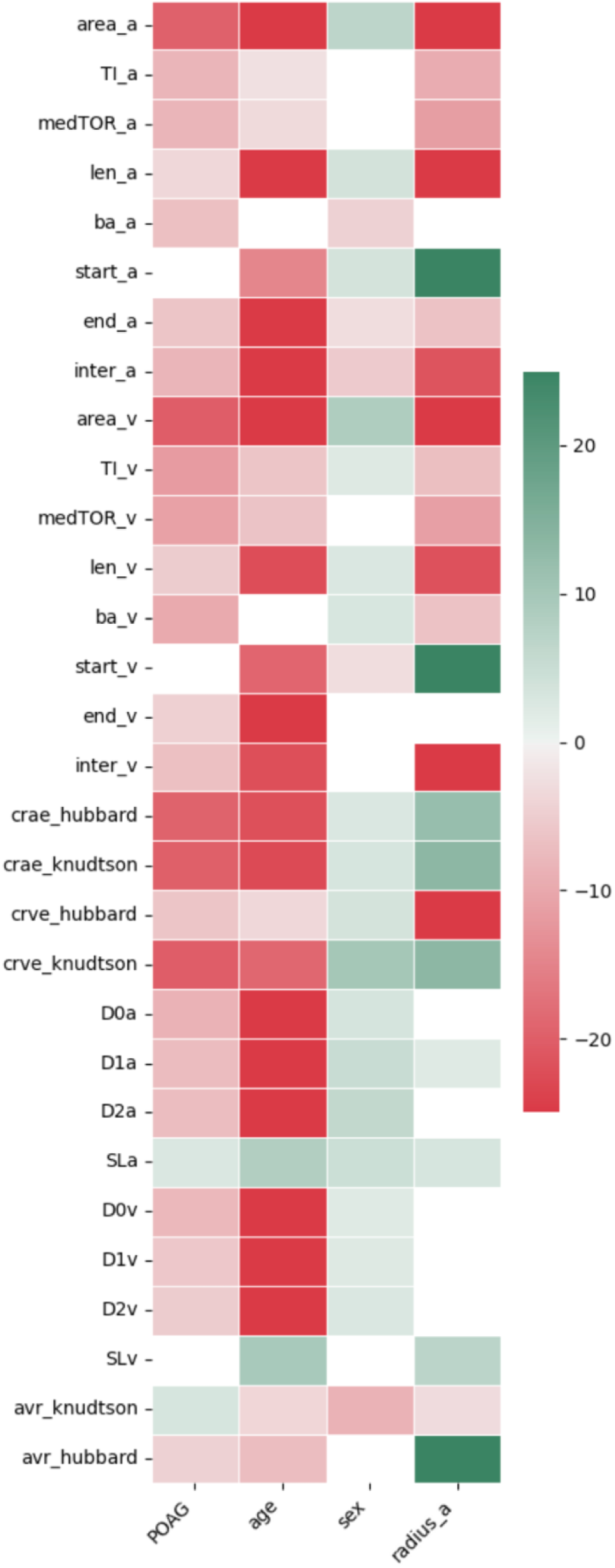
Results of the statistical analysis. Non-significant p-values are in white. Significant p-values are represented by Z values in the heatmap. Z values larger than 25 or smaller than −25 are clipped to these color values.

**Table 3:**
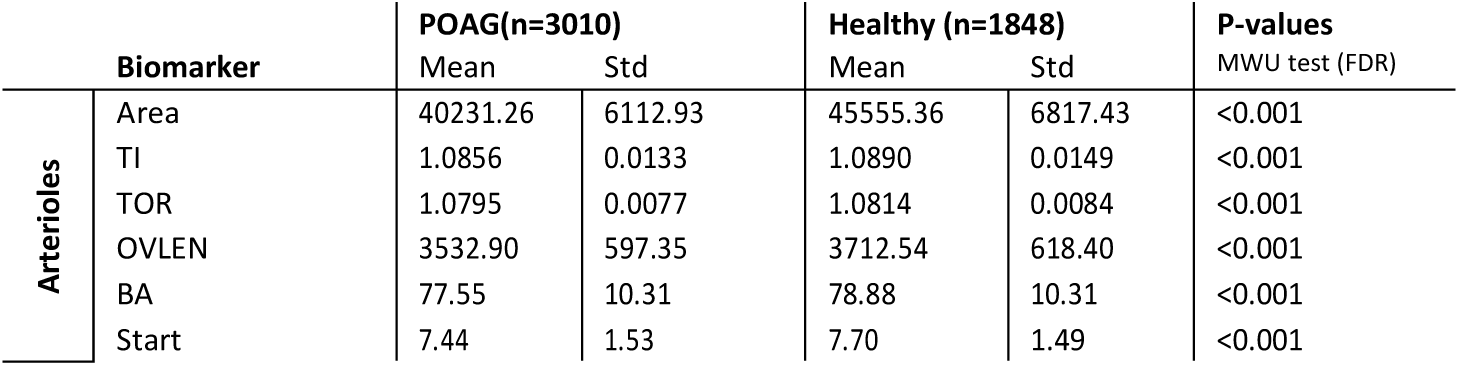
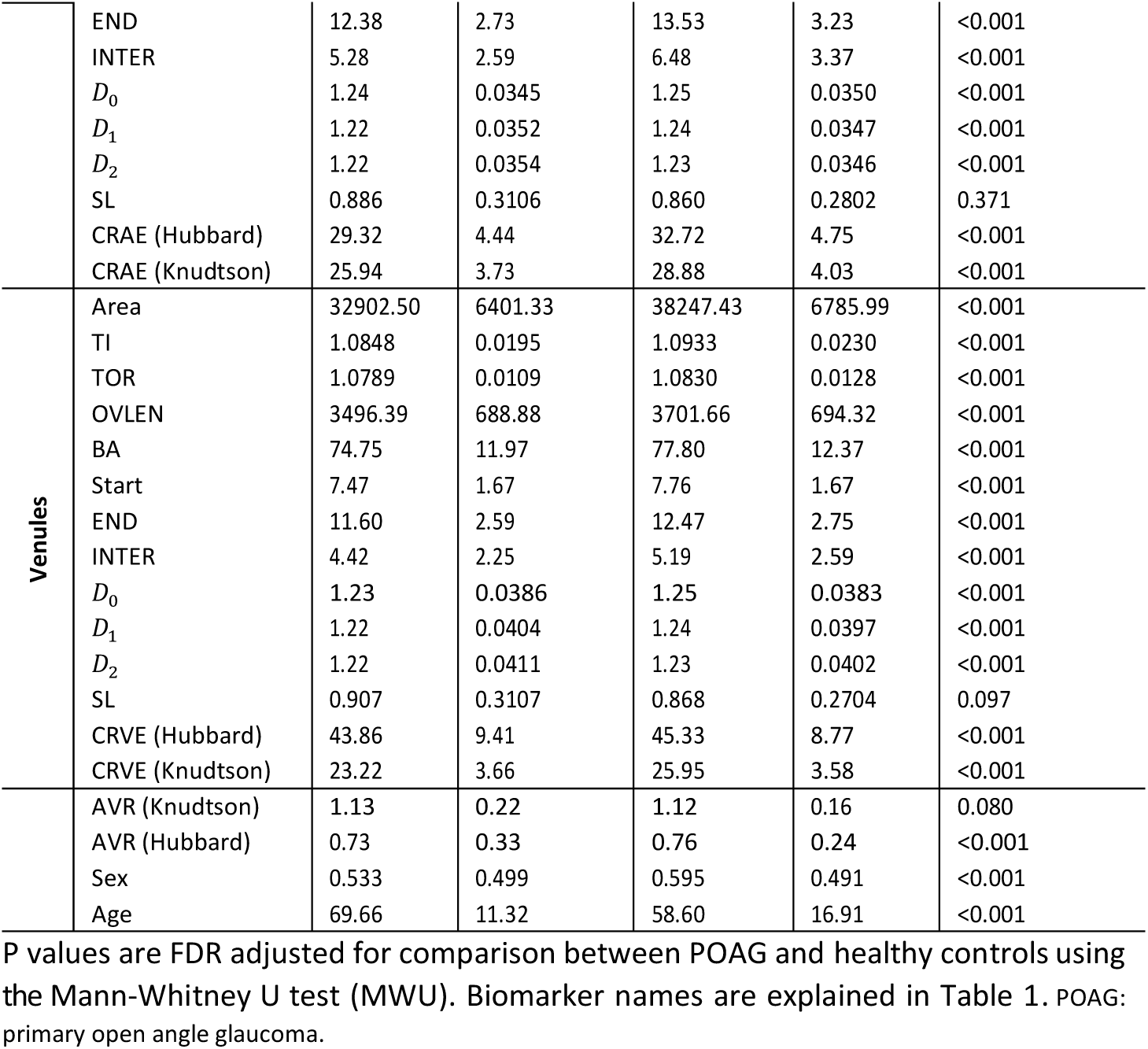
Summary of the computed VBMs using their implementation from PVBM.

A boxplot illustrating the arteriolar area, venular tortuosity, CRAE, and CRVE (according to Knudtson’s method) segmented by diagnosis and sex relative to patient age is depicted in Fig. 5. Additional boxplots covering other vascular biomarkers are available in the Appendix, as shown in Fig. A3. These boxplots show a trend in the VBMs with respect to the age, gender, and POAG diagnosis illustrating the outcome of the statistical analysis.

**Fig. 5:**
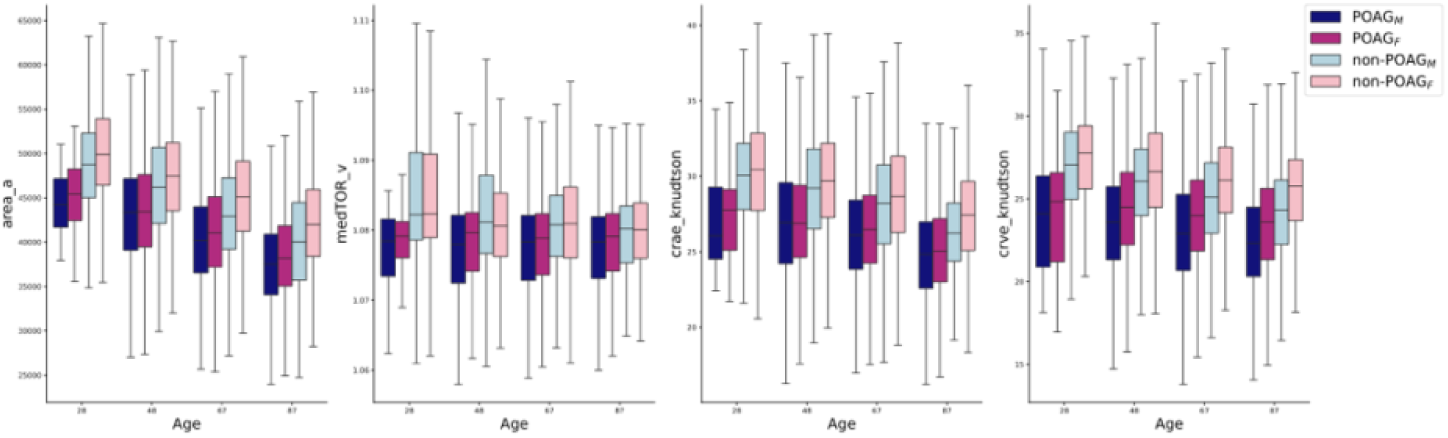
Boxplot of the Area*_a_*, TOR*_v_*, CRAE and CRVE (Knudtson), grouped by diagnosis and gender with respect to the patient age.

## Discussion

In this work, we propose an end-to-end pipeline for the automated analysis of the retinal vascular geometry in a large population cohort, for which POAG was used as a case example. Fully automatization differs from previous work in which the VBM computation was semi-supervised^1–3, 9, 14–19, 40^.

All computed retinal vascular parameters are lower in participants of higher age, concordant with pre-existing literature^14, 21, 41, 42^. Women have slightly higher arteriolar and venular areas (Area_a_), due to longer and wider vessels (OVLEN_a_), and slightly higher arteriolar and venular fractal dimension levels. This is to our knowledge the first time significant sex differences are reported for these biomarkers. The Beaver Dam Eye study^42^ found narrower CRAE and CRVE in men and the Multi-Ethnic Study of Atherosclerosis^41^ narrower CRAE in men, which we replicated. Incident cardiovascular comorbidity (e.g. hypertension prevalence) between the sexes is not reported in these studies, as in the current work, and could represent a potential bias.

The work by Zhu et al. is a recent example of the fact that age can be estimated from DFIs using a deep learning model (correlation of 0.81, p*<*0,001)^43^. Similarly, previous work showed that sex can be identified from DFI^44^. For these tasks, end-to-end deep learning models are used and the gain in performance is reached at the expense of explainability. The automated approach presented in this work at least partly tackles the explainability limitation of end-to-end deep learning models e.g. age and sex estimation characterizing age- and sex-related retinal vascular changes.

Despite extensive research efforts^45, 46^, the precise mechanism underlying POAG remains elusive. The mechanical theory posits that mechanical stress induces damage to the ganglion cells and their axons. Factors such as corneal thickness, corneal hysteresis, and the pressure gradient across the lamina cribrosa contribute to the susceptibility of the optic nerve to fluctuations in IOP, resulting in axonal injury along the optic nerve^47–49^. Alternatively, possibly complimentary, the vascular theorem suggests that optic neuropathy arises from repetitive ischemic insults or subtle changes in perfusion^50–52^. The increased prevalence of normal tension glaucoma (NTG, POAG subcategory with a maximal untreated IOP below 21 mmHg) in individuals with conditions like migraine, Raynaud’s disease, and Flammer syndrome supports this hypothesis^53, 54^. Additionally, the independent association between elevated arterial blood pressure variability and extremes of blood pressure levels, and the incidence and progression of POAG, alongside the elevated prevalence of cardio/reno/neurovascular disease (hyperlipidemia, metabolic syndrome, ischemic heart disease, stroke small vessel disease, chronic kidney disease) in POAG patients, provide further support for the interrelation between POAG and systemic vascular dysregulation^55–58^. This work is the first to explore the retinal vascular features of glaucoma patients on a large scale, thus facilitating a more robust interpretation and statistical validity of vascular theory.

Several studies indicated significantly narrower CRAE and CRVE in POAG patients, compared to controls without POAG^4,6, 10–13, 59^. Furthermore, a decrease in CRAE was previously associated with an increased incidence of POAG after 10 years of follow-up and progression in patients with NTG after 2 years of follow-up^5, 9^. In patients with asymmetric disease, CRAE and CRVE were narrower in the eye with more severe disease^8^. In bilateral glaucoma suspects who converted to unilateral glaucoma, a narrower CRAE is found at baseline and both narrower CRAE and CRVE are found at the time of conversion, compared to non-converted eyes. CRAE and CRVE were significantly narrower at the time of conversion than at baseline, at least in converted eyes^7^. Yoo et al. showed that CRAE had the largest area under the ROC-curve, compared to CRVE and AVR, for discriminating between POAG patients and healthy eyes, showing no significant difference with AUC of retinal nerve fiber layer (RNFL) thickness^12^. On the other hand, several studies, including the Rotterdam Study and the Beaver Dam Eye Study, report an insignificant association between CRAE/CRVE and POAG^42, 60–62^. Furthermore, several more studies, including the Blue Mountains Eye study, report a non-significant relationship between CRVE and POAG prevalence^4, 5, 9, 11, 12^. Of note, all these studies consistently had relatively low sample sizes, mostly under 100 POAG cases, never surpassing more than 250 cases, and exhibit clear heterogeneity in vessel segmentation method and quality of retinal images. This work, comparing multiple images of more than 3000 POAG patients and more than 1800 controls and a high level of standardization and quality control, can be seen as a definitive conclusion that both CRAE and CRVE are on average lower in POAG patients after adjustment for age, sex and disc radius a. Consequently, AVR did not significantly differ between POAG patients and controls, in line with a lowering of both CRAE and CRVE.

Tortuosity, branching angle, and fractal dimension reflect the circulatory optimality of the retinal microvasculature, optimizing blood flow with minimal energy expenditure^15,63^. Tortuosity is associated with tissue hypoxia, potentially mediated by secretions from vascular endothelial cells, which regulate blood flow through mediators such as nitric oxide and endothelin, which influence angiogenesis and therefore vessel geometry^64–66^. Lower tortuosity has been associated with aging^67^. Some studies, including the EPIC-Norfolk Eye Study, did not show a significant association between arteriolar and venular tortuosity in POAG patients compared to controls^11, 13, 61^. However, straighter and less tortuous arterioles and venules were significantly associated with POAG and NTG separately in other trials and are confirmed in this work^21, 60^. In the literature, the branching angle was found significantly different on the arteriolar or venular side in POAG patients compared to healthy controls^21, 60^. Increased branching angles have been related to decreased blood flow, whereas decreased angles have been related to ageing and hypertension^68, 69^. This study confirms both the lower arteriolar and venular branching angles in POAG patients. The lower fractal dimension, both arteriolar and venular, as measured with optical coherence tomography angiography, was previously observed in POAG patients compared to controls and is confirmed in this work^13,21, 70^.

In conclusion, after adjustment of both age, sex and disc radius a, POAG patients seem to exhibit lower arteriolar and venular diameter, area, tortuosity, branching angle, endpoints, intersection points and fractal dimension levels. These changes mimic those seen in higher age categories, which underlines the possibility of pronounced vascular aging and dysfunctional vascular endothelium in POAG patients.

## Limitations and future directions

The vascular effects shown in this paper do not prove the vascular theorem but point to a certain association. Similarly, Tham et al.^10^ observed no association between CRAE and POAG prevalence after adjusting for RNFL thickness, in contrast with CRVE - which showed a significant association after adjustment. For both CRAE and CRVE, the indirect effects, being the associations mediated through RNFL thickness, were significant and were 32.3% and 12.3% of the total effect respectively. In analogy, this association cannot necessarily be used in favor of the vascular theorem as the vascular changes can occur secondary to preceding RNFL loss due to other reasons. Longitudinal analyzes of retinal vascular changes and RNFL thinning in glaucoma progression has the potential to further elucidate this discussion and represent the subject of future research. The disc size did influence the parameters. As the disc radius increased, vessel area and length, calculated in a fixed ROI excluding the OD, logically decreased and the number of starting points increased. Both CRAE and CRVE, calculated in zone B that depends on the disc radius a, tended to increase in higher radii. These differences point to the need for adjustment of the disc size in automated VBM pipelines.

This is the first time a fully automated end-to-end pipeline is published for the analysis of retinal vascular geometry in a large cohort (13,185 patients). Both arteriolar and venular diameter, area, length, tortuosity, branching angle, end-points, intersection points and fractal dimension levels are independently lower in POAG patients and older patients, after adjustment for sex and disc radius a. Given the independent similarities in retinal vascular geometry changes between older age and POAG prevalence the theory of pronounced vascular ageing in POAG patients is proposed.

## Data Availability

All data produced are available online at https://pvbm.readthedocs.io/en/latest/

https://pvbm.readthedocs.io/en/latest/

## Authors contribution

JB, JVE, IS conceived and designed the research, JF developed the algorithms and performed the analysis under the supervision of JB. LB checked the statistical analyses. JVE, IS, OA contributed and curated the dataset. JVE, IS provided the physiological interpretation of the results and wrote the discussion section. ARB, JVE guided the statistical analysis. JB and JF drafted the first version of the manuscript, JF, LB prepared the figures. All authors edited and revised the manuscript and approved the final version.

## Acknowledgment

We acknowledge the assistance of ChatGPT, an AI-based language model developed by OpenAI, for its help in editing the English language of this manuscript.

## Abbreviations and Acronyms

A: arterioles
CRAE: central retinal artery equivalent
CRVE: central retinal vein equivalent
DFI: digital fundus images
HTG: high-tension glaucoma
LMM: linear mixed-effects model
NTG: normal tension glaucoma
OD: optic disc
POAG: primary open-angle glaucoma
ROI: region of interest
V: venules
VBMs: vascular biomarkers

## List of Additional Figures

**Fig. A1:**
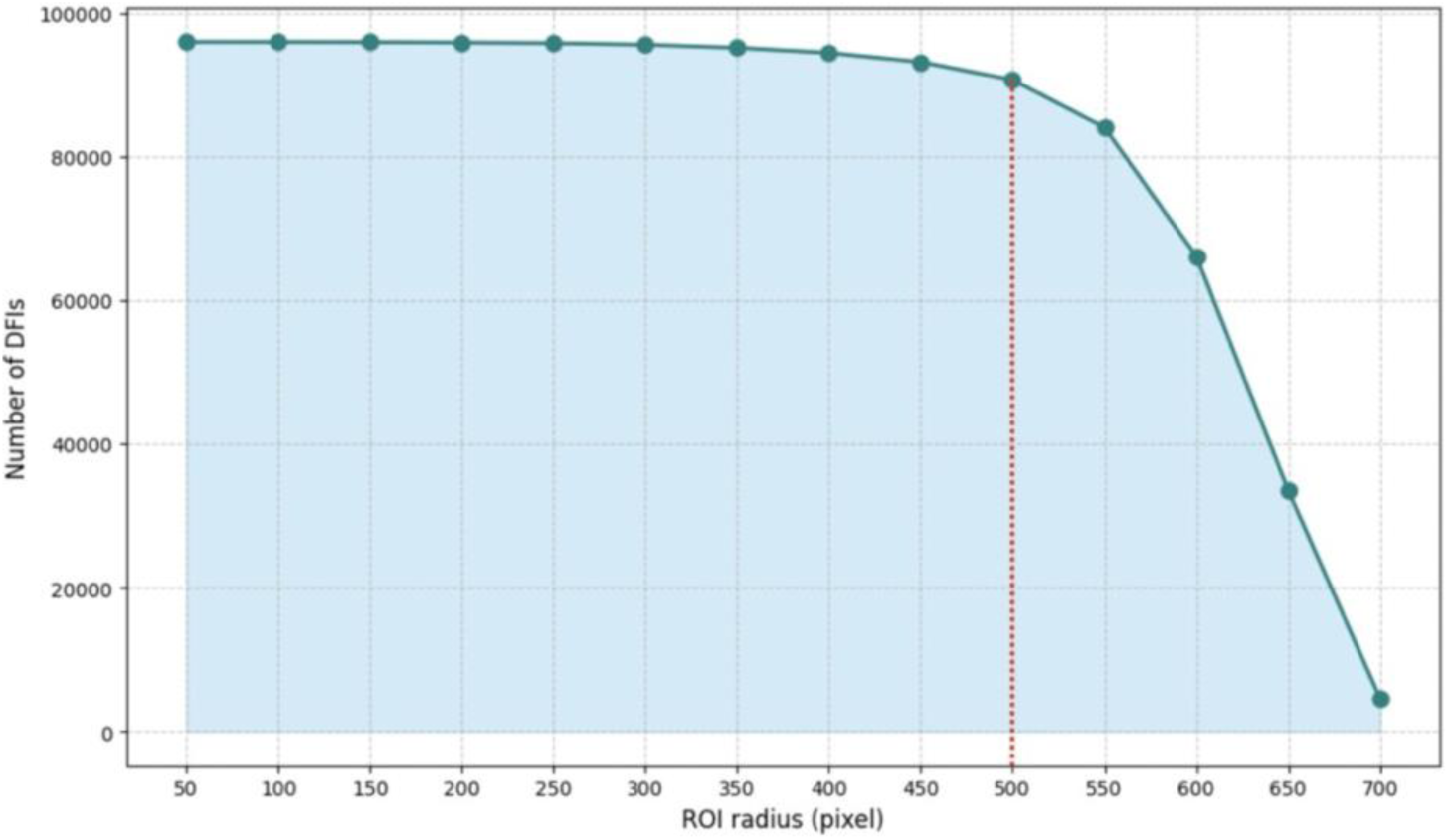
Number of DFIs included in the analysis with respect to the size of the ROI.

**Fig. A2:**
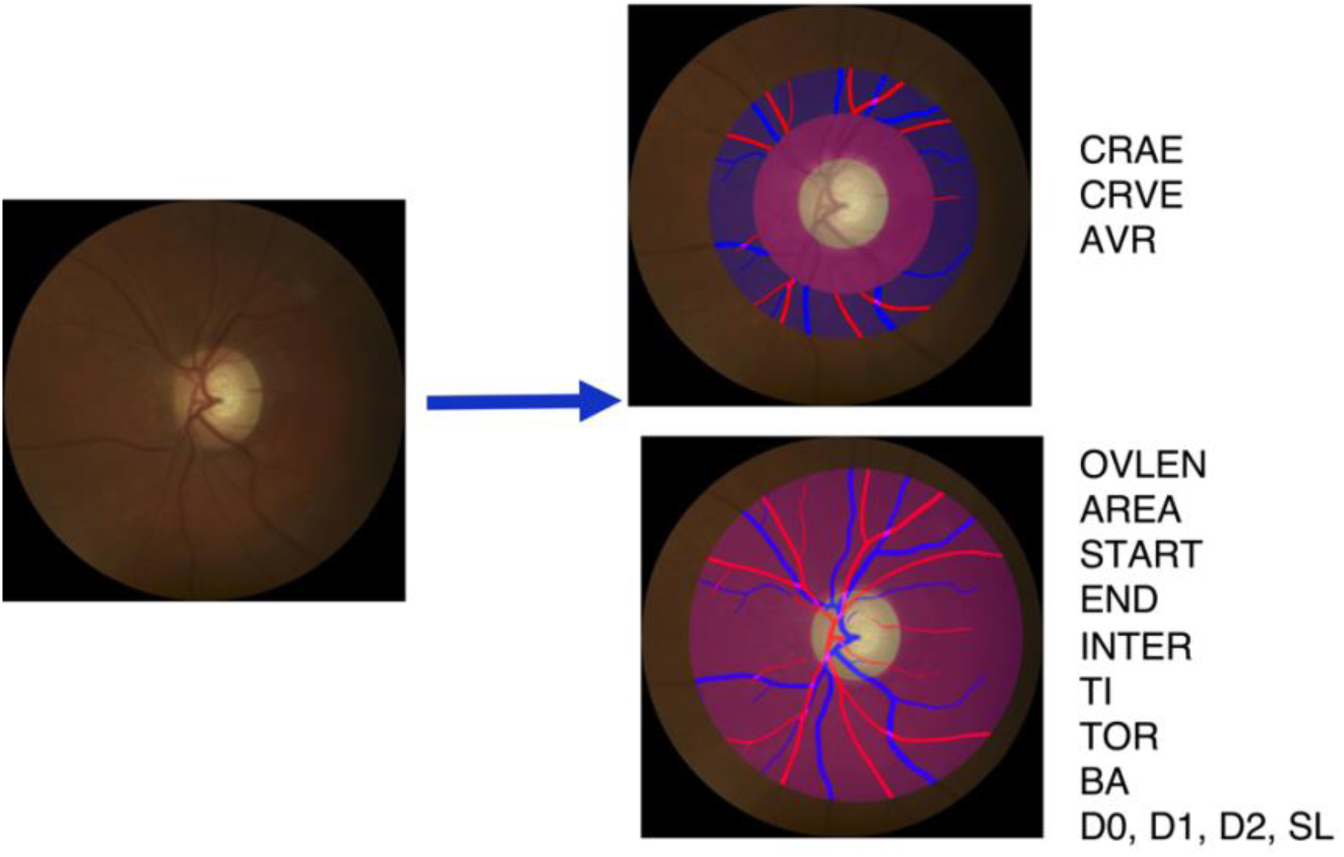
Overview of a DFI segmentation in the zone B and fixed size ROI.

**Fig. A3:**
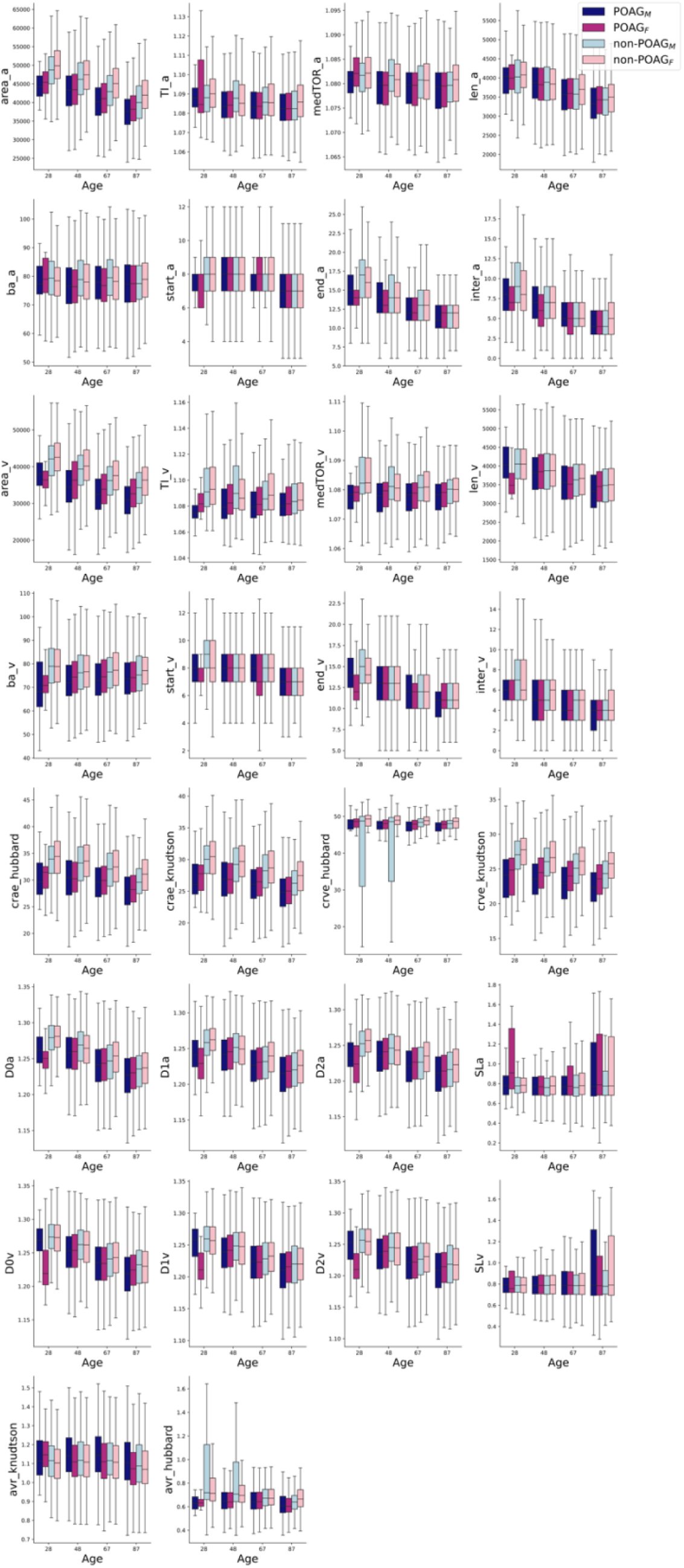
Boxplots for each VBM, grouped by diagnosis and gender with respect to the patient age.

